# Heart Disease Deaths during the Covid-19 Pandemic

**DOI:** 10.1101/2020.06.04.20122317

**Authors:** Jeremy S. Faust, Zhenqiu Lin, Harlan M. Krumholz

## Abstract

The SARS-CoV-2 pandemic is associated with a reduction in hospitalization for an acute cardiovascular conditions. In a major health system in Massachusetts, there was a 43% reduction in these types of hospitalizations in March 2020 compared with March 2019.4 Whether mortality rates from heart disease have changed over this period is unknown.

We assembled information from the National Center for Health Statistics (Centers for Disease Control and Prevention) for 118,356,533 person-weeks from Week 1 (ending January 4) through Week 17 (ending April 25) of 2020 for the state of Massachusetts. We found that heart disease deaths are unchanged during the Covid-19 pandemic period as compared to the corresponding period of 2019. This is despite reports that admissions for acute myocardial infarction have fallen during this time.

---

The Covid-19 pandemic has been associated with a reduction in hospitalization for^1^,^2^,^3^,^4^ In a major health system in Massachusetts, there was a 43% reduction in these types of hospitalizations in March 2020 compared with March 2019.^4^ Whether mortality rates from heart disease have changed over this period is unknown.

Accordingly, we assembled data on weekly heart disease deaths in Massachusetts from the National Center for Health Statistics (Centers for Disease Control and Prevention) for 118,356,533 person-weeks from Week 1 (ending January 4) through Week 17 (ending April 25) of 2020.^5^ A total of 4,085 heart disease deaths were recorded during this period (incidence rate, 3.5 per 100,000 person-weeks; 95% confidence interval [CI] 3.4 to 3.6), versus 4,209 heart disease deaths over the same period in 2019 (incidence rate, 3.6 per 100,000 person-weeks; 95% CI, 3.5 to 3.7) (incidence rate ratio, 0.97; 95% CI, 0.92 to 1.01) (Figure 1). Covid-19 deaths and overall deaths from all causes, for comparison, markedly rose over the study period.

**Figure 1.**
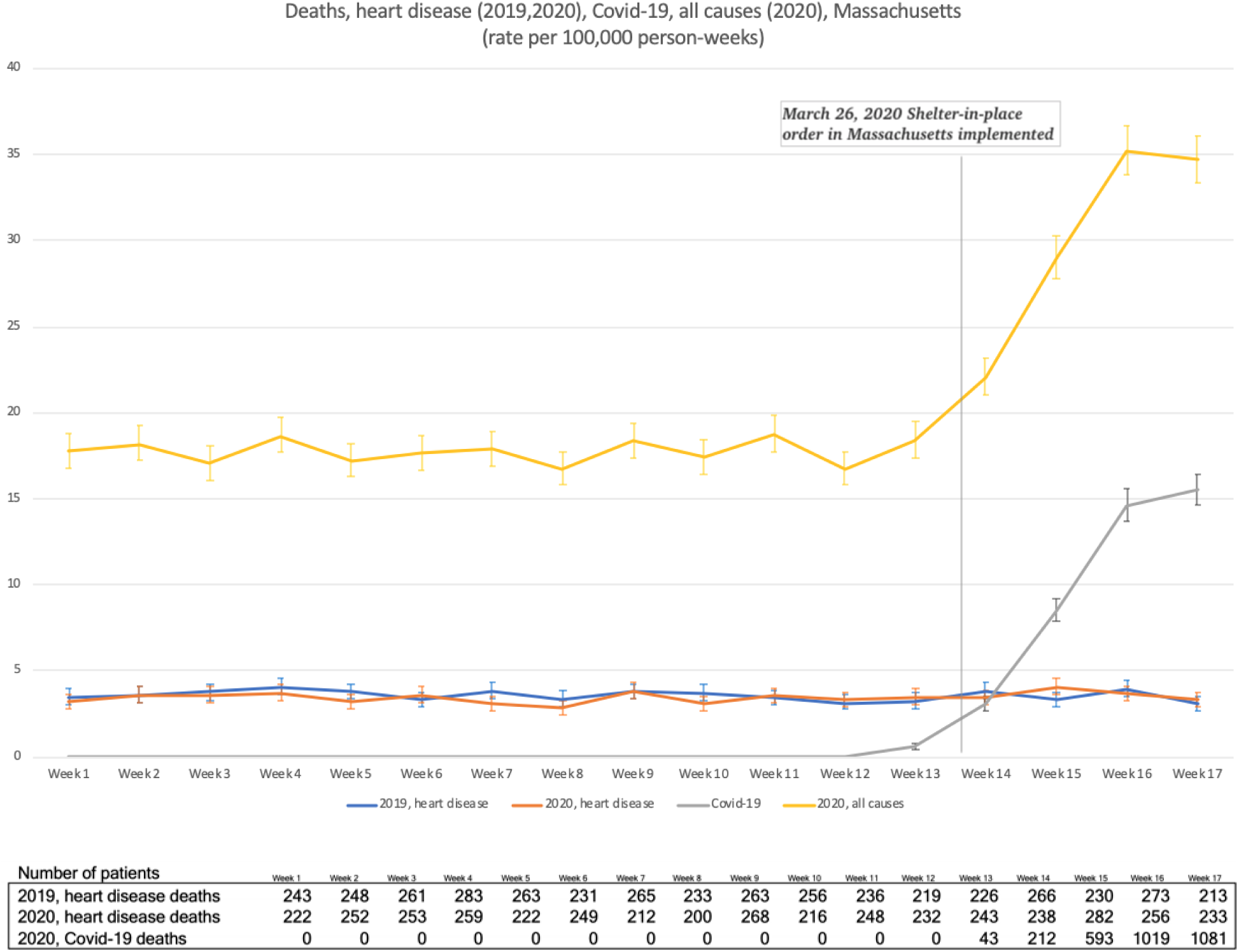
Incidence of Heart Disease deaths before and during the Covid-19 Pandemic in 2020 and during the Same Period in 2019, Relative to the Incidence of deaths from Covid-19 and All-Cause Deaths during 2020. Shown are data from Massachusetts. The data shown in orange are the reported weekly incidence rates of heart disease per 100,000 person-weeks during the period from Week 1 through Week 17, 2020. The data shown in blue are the reported weekly incidence rates of heart disease per 100,000 person-weeks during the period from Week 1 through Week 17, 2019. 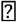 bars indicate 95% confidence intervals. The data shown in gray are the rates for Covid-19 deaths per 100,000 person-weeks for the same period, with the first Massachusetts death reported March 26, 2020. The data shown in yellow are the numbers of all-cause deaths per 100,000 person-weeks.

There are several feasible explanations for these findings. It is possible that some cardiovascular deaths could be incorrectly attributed and coded as Covid-19 deaths, though there is not routine post-mortem SARS-CoV-2 testing of patients who died in non-medical settings, nor instructions to medical officials to presume deaths of unknown cause to be coded as Covid-19-related in Massachusetts. The observed >40% decline in hospitalizations, while large, may not be associated with a decline in cardiovascular deaths, owing to the relatively low number of related modifiable deaths during the short time period of interest. There could be an actual decline in acute cardiovascular emergencies as a result of changes in exposures and behaviors during the Covid-19 shutdown. Finally, the reporting of cardiovascular deaths may not yet be sufficiently complete for 2020. However, the CDC reports that the currently available figures are estimated to be greater than 98.3% to 99.3% complete for the weeks reported here, and the final 0.7% to 1.7% is unlikely to change the qualitative conclusions (Supplementary Appendix, Table S1).^6^

These findings add more information to the pursuit of understanding the change in cardiovascular healthcare utilization during the Covid-19 outbreak in the United States but are unable to identify any definitive cause. For now, the information is at least reassuring that there was not a marked increase in cardiovascular deaths in Massachusetts during the early phase of the outbreak, despite a decrease in the number of patients seeking acute care for related emergencies. acute cardiovascular conditions.

## Data Availability

The data is available to the public.
https://www.cdc.gov/nchs/data/vsrr/report001.pdf

